# Infection Control of 2019 Novel Corona Virus Disease (COVID-19) in Cancer Patients undergoing Radiotherapy in Wuhan

**DOI:** 10.1101/2020.03.21.20037051

**Authors:** Conghua Xie, Xiaoyong Wang, Hui Liu, Zhirong Bao, Jing Yu, Yahua Zhong, Melvin L.K. Chua

## Abstract

**Background:** A pandemic of 2019 novel corona virus disease (COVID-19), which was first reported in Wuhan city, has affected more than 100,000 patients worldwide. Patients with cancer are at a higher risk of COVID-19, but currently, there is no guidance on the management of cancer patients during this outbreak. Here, we report the infection control measures and early outcomes of patients who received radiotherapy (RT) at a tertiary cancer centre in Wuhan.

**Methods:** We reviewed all patients who were treated at the Zhongnan Hospital of Wuhan University (ZHWU) from Jan 20 to Mar 6, 2020. This preceded the city lock-down date of Jan 23, 2020. Infection control measures were implemented, which included a clinical pathway for managing suspect COVID-19 cases, on-site screening, modifications to the RT facility, and protection of healthcare workers. Primary end-point was infection rate among patients and healthcare staff. Diagnosis of COVID-19 was based on the 5^th^ edition criteria.

**Findings:** 209 patients completed RT during the study period. Median age was 55 y (IQR = 48-64). Thoracic, head and neck, and lower gastrointestinal and gynaecological cancer patients consisted the majority of patients. Treatment sites included thoracic (38.3%), head and neck (25.4%), and abdomen and pelvis (25.8%); 47.4%, 27.3%, and 25.4% of treatments were for adjuvant, radical, and palliative indications, respectively. 188 treatments/day were performed prior to the lock-down, in contrast to 12.4 treatments/day post-lock-down. Only one (0.48%) patient was diagnosed with COVID-19 during the study period. No healthcare worker was infected.

**Interpretation:** Herein, we show that in a susceptible population to COVID-19, strict infection control measures can curb human-to-human transmission, and ensure timely delivery of RT to cancer patients.

**Funding:** This study was funded by Health Commission of Hubei Province Scientific Research Project, WJ2019H002, Health Commission of Hubei Province Medical Leading Talent Project.

**Research in context:** *Evidence before this study:* The 2019 novel coronavirus disease (COVID-19) is now a global pandemic. Cancer patients are at risk of COVID-19 pneumonia, and thus infection control measures are crucial to mitigate their risk of infection. We searched PubMed and Medline for articles published up to Mar 12, 2020, using the following keywords: “COVID-19”, “SARS”, “SARS-CoV-2”, “infection control”, and “cancer”. No evidence exists that informs on the appropriate infection control measures for COVID-19.

*Added value of this study:* We report our single centre experience on the detailed infection control measures that were undertaken to minimise cross transmission between cancer patients undergoing radiotherapy, and between patients and healthcare workers. Measures entailing screening of suspect cases, re-organisation of the treatment facility, and protection of healthcare workers were described. With our infection control protocol, we recorded only one COVID-19 case among the 209 patients (0.48%) who were treated at our centre during the period of Jan 20 to Mar 6, 2020. No healthcare worker was affected.

*Implications of all the available evidence:* The effective infection control measures outlined in this study will help institutions worldwide affected by COVID-19 to formulate guidelines to mitigate nosocomial human-to-human transmission, especially among susceptible patients.

## Introduction

Globally, there is an unprecedented outbreak of acute respiratory illness, named 2019 novel coronavirus disease (COVID-19), which is caused by a severe adult respiratory syndrome coronavirus 2 (SARS-CoV-2).^1,2^ Wuhan was the virus epicentre where SARS-CoV-2 first emerged, but rapidly, this has become a pandemic affecting nearly 90 countries worldwide. As of Mar 12, 2020, a total of 80,981 confirmed cases and 3,173 death cases have been reported in China, of which an overwhelming majority was from Wuhan city and Hubei province. Due to the rapid human-to-human transmission and the potential severity of COVID-19, hospitals from the outbreak areas have been severly affected by the overload of infected patients, which has led to the suspension of several healthcare services.

For cancer patients, a delay of diagnosis and treatment can be detrimental to survival.^3^ However, cancer patients are also at risk of SAR-CoV-2 infection, as reported by two case series from China, which may be attributed to immunosuppressive treatments and recurrent visits to the hospital.^4,5^ Thus, robust infection control measures are necessary to mitigate the risk of contracting COVID-19 in cancer patients.

Radiotherapy (RT) constitutes a primary modality for the treatment of cancer, with approximately 50% of all cancer patients requiring RT during their cancer journey.^6^ Often, it is used as a definitive or adjuvant modality, and in patients with disseminated systemic disease, RT is effective for palliating symptoms. It is therefore imperative that RT services are not affected during this outbreak. Here, we report our experience and preliminary outcomes of 209 RT patients, who were treated at the Zhongnan Hospital of Wuhan University (ZHWU) during the period when the city was locked down on Jan 23, 2020.

## Methods

### Patient cohort

All patients who were treated at the Department of Medical and Radiation Oncology, ZHWU, from Jan 20 to Mar 6, 2020 were included in this study. The start-date of the study period preceded the commencement of the lock-down of Wuhan city on Jan 23, 2020. Patient’s demographics, medical history, contact history, clinical symptoms and examination, haematological and biochemical investigations, and results of the computed tomography (CT) of the chest were obtained. Data cut-off date for the study was Mar 6, 2020. This study was approved by the institutional review board of ZHWU (2020039). As aggregated, anonymised patient data was used in this study, waiver of consent was approved by the ethics committee.

### Infection control measures

The workflow for screening of both hospitalised and non-hospitalised patients is outlined in **Figure 1**. Identification of suspect cases with COVID-19 infection was the first crucial step in infection control. In this regard, routine pre-screening of the patient and family members should be performed. Daily screening of a constellation of clinical symptoms included: 1) body temperature of ≥37.3°C; 2) presence of fatigue, respiratory symptoms, or gastrointestinal symptoms (diarrhea, abdominal pain etc.); and 3) a contact history with SARS-CoV-2 infected patients. If patients recorded any of the above symptoms, we would refer them to a comprehensive set of investigations, including a CT chest using our departmental scanner (Sensation Cardiac 64x, Siemens, Germany) and laboratory work-up, comprising of a complete blood count and biochemical analyses (coagulation profile, hepatic and renal function, erythrocyte sedimentation rate, C-reactive protein, procalcitonin, lactate dehydrogenase, and creatine kinase). Of note, the characteristic CT change of COVID-19 pneumonia is the presence of ground-glass changes in the outer zone of one or both lungs. This method of diagnosis for COVID-19 is considered more reliable than the SARS-CoV-2 real-time reverse transcription polymerase chain reaction (RT-PCR) assay, since CT manifestations tend to appear earlier than nucleic acid detection.^7^

**Figure 1.**
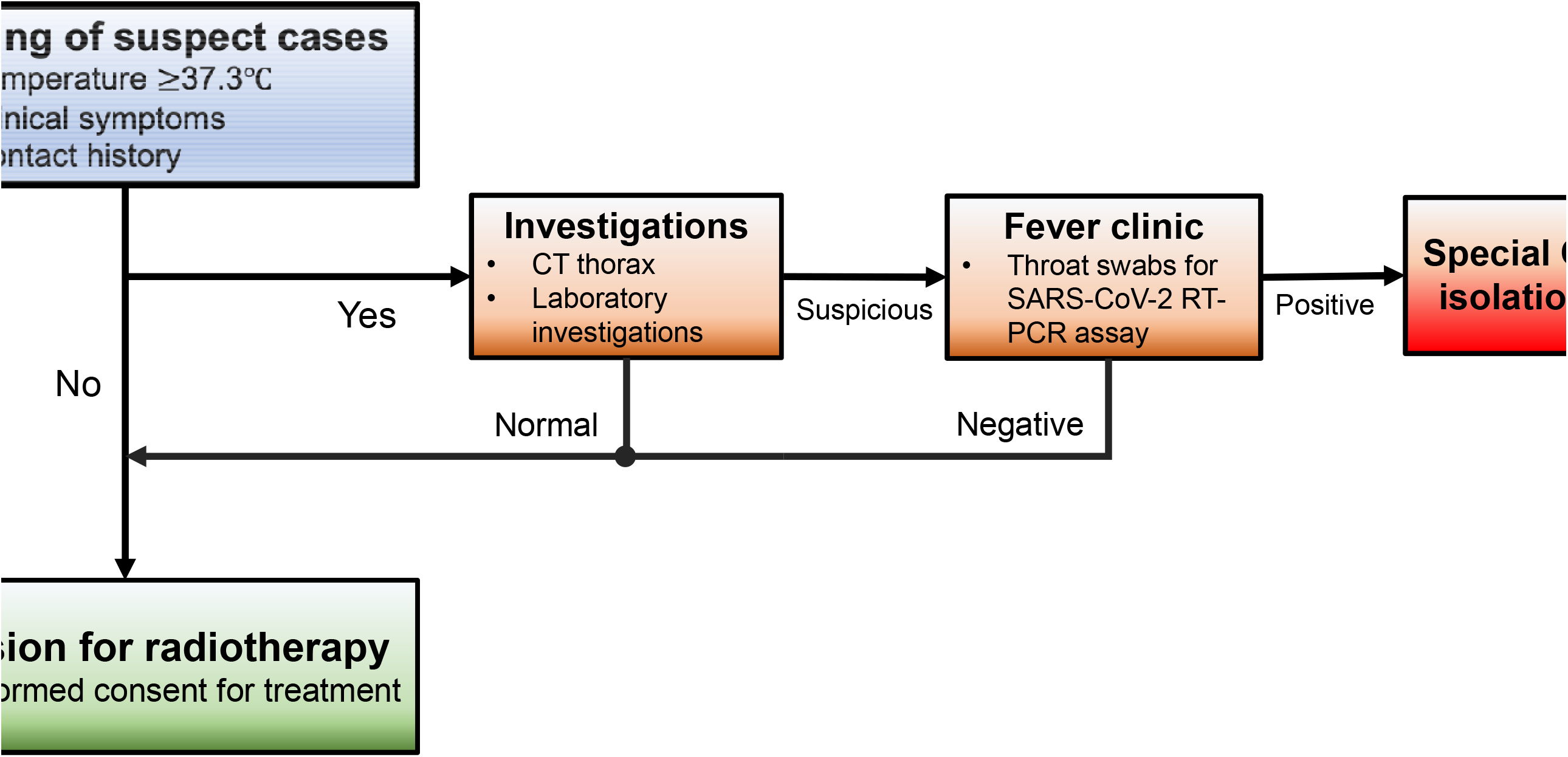
Clinical pathway for screening of suspect COVID-19 cases for patients with cancer undergoing radiotherapy. A three-step approach was adopted to escalate investigations for suspect cases. We established a fever clinic to consult patients with a suspicious CT and laboratory investigations. Patients with a positive COVID-19 diagnosis, ascertained by either abnormal CT or RT-PCR test will be transferred to a dedicated facility for isolation and treatment. Abbreviations: CT, computed tomography; SARS-CoV-2, severe adult respiratory syndrome novel coronavirus 2; RT-PCR, real-time reverse transcripton polymerase chain reaction; COVID-19, 2019 novel coronavirus disease.

If either the CT or laboratory results were abnormal, the suspect cases would then be sent to the outpatient fever clinic for further assessment. Throat swab samples would be collected and tested using the SARS-CoV-2 RT-PCR assay. While waiting for the outcome of the RT-PCR test, patients would be quarantined in a dedicated area within the hospital compound to mitigate the risk of nosocomial spread. Contact, droplet and airborne precautions were undertaken for the suspect cases. In the event of a positive RT-PCR result or highly suspicious CT findings, patients would be transported to a designated infectious disease hospital for isolation, observation, and treatment. For patients with normal results, they will be admitted to the hospital for RT or receive RT as an outpatient.

### On-site screening

We employed the following workflow to screen all patients and health-care workers (nurses, radiation therapists, physicists, physicians and paramedics): 1) All people entering the treatment area must record a body temperature of less than 37.3°C prior to entry; 2) they would be screened again for contact history; 3) they must don a three-ply surgical mask at all times; and 4) strict hand hygiene must be practised.

### Infection control measures at the RT treatment unit

Patient holding area should be well-ventilated. Central air conditioning in the treatment area was not permitted. The treatment devices in close contact with the patient were thoroughly disinfected after every patient, while patient-specific treatment gadgets (e.g. thermoplastic shells, VacLoks etc.) were returned to the respective position after each treatment to avoid contact contamination.

Next, to avoid overcrowding, treatment timings were staggered such that number of patients was limited to two for every 30-40 min time-slot. Strict distancing of at least 1.5 m apart was ensured, and patients were not allowed to communicate with each another. Finally, daily disinfection measures (cleaning and sterilisation of all surfaces, including seats, tables, floors and walls of the waiting hall) were performed for the entire treatment facility. The detailed patient flow and floor plan of the treatment facility is illustrated in **Figure 2**.

**Figure 2.**
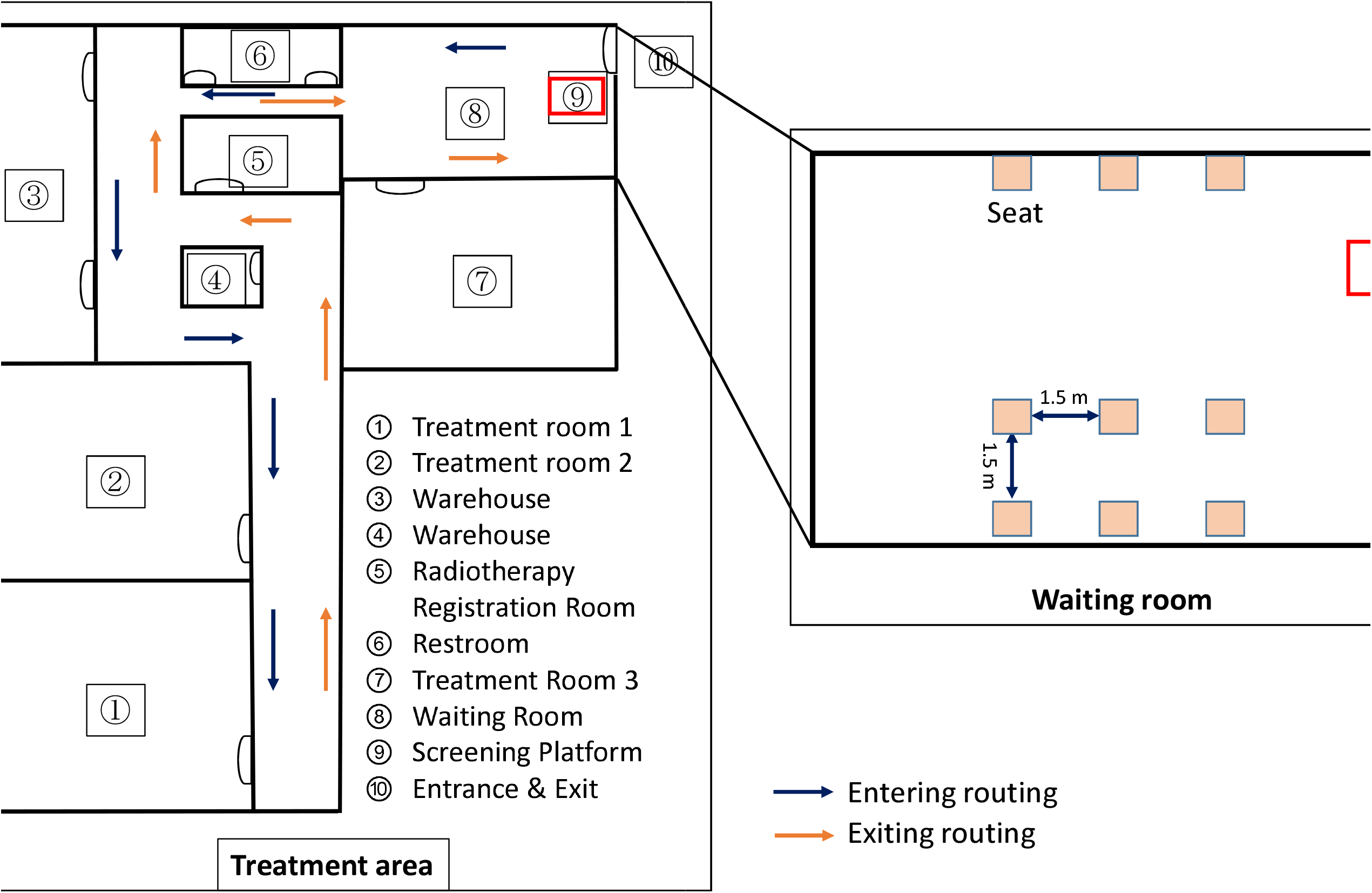
Layout of the radiotherapy centre at the Zhongnan Hospital of Wuhan University. The facility was re-orgnised to accommodate an on-site screening platform and a spacious patient holding area. Distancing between patients was ensured to minimise contact. We also dermacated fixed entry and exit points for patient flow through the radiotherapy facility. Floor plan not drawn to scale.

### Protection of health-care workers

Given the risk of COVID-19 pneumonia to frontline medical staff, the following measures were implemented to mitigate the risk of cross-infection from patients to healthcare workers: 1) a buddy system to expedite the notification of unwell team members who harboured symptoms of fever, fatigue and dry cough, or who had a positive contact history with a COVID-19 patient; 2) twice daily temperature monitoring of all staff; 3) all staff members who were involved in transfer and delivery of treatment for patients had to don protective gear comprising of disposable surgical caps, gowns, N95 masks, face screens, double-layered gloves, and shoe covers; 4) radiation therapists were split into two-man teams at any point in time, along with optimisation of processes for patient set-up and treatment delivery to reduce cross infection; 5) protective gear should be disposed of in a medical hazard waste bag; 6) strict hand hygiene before leaving the treatment area; 7) team gatherings during work were prohibited; and 8) finally, social distancing was encouraged to prevent transmission to family members.

### Outcomes

Primary outcome of this study was infection rate among patients and healthcare staff. Diagnosis of COVID-19 was based on the 5^th^ edition criteria (see **Supplementary appendix**). Clinical and treatment parameters, including RT indications, number of RT sessions per day, and treatment delays were analysed. Survival status of all patients were updated as of Mar 12, 2020.

## Results

### Patient characteristics

From Jan 20, 2020 to Mar 6, 2020, 209 patients were treated at our RT facility. **Table 1** summarises the clinical characteristics of patients and their treatment details. Median age of patients was 55 y (IQR = 48-64). Gender was evenly distributed. Thoracic cancers (N = 80 [38.3%], including lung, breast, and oesophageal cancers), head and neck cancers (N = 53 [25.4%]), and lower gastrointestinal and gynaecological cancers (N = 54 [25.8%]) comprised the majority of the study cohort. Of these, 172 (82.3%) were inpatients, while 37 patients (17.7%) communted daily for treatment.

**Table 1.**
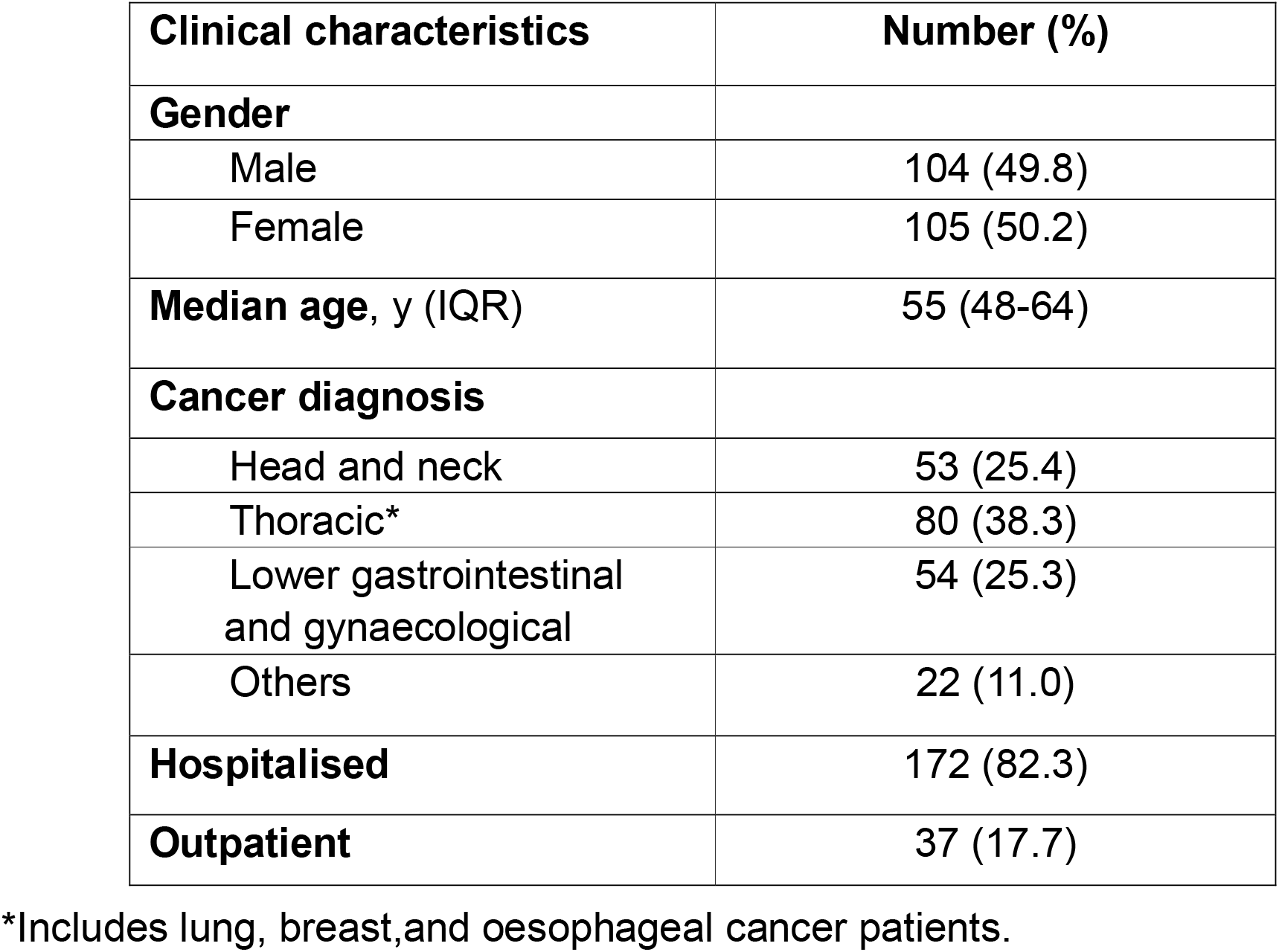
Characteristics of 209 cancer patients treated at Zhongnan Hospital of Wuhan University (ZHWU) during the COVID-19 epidemic.

| **Clinical characteristics** | **Number (%)** |
| --- | --- |
| **Gender** |  |
| Male | 104 (49.8) |
| Female | 105 (50.2) |
| **Median age, y (IQR)** | 55 (48-64) |
| **Cancer diagnosis** |  |
| Head and neck | 53 (25.4) |
| Thoracic* | 80 (38.3) |
| Lower gastrointestinal and gynaecological | 54 (25.3) |
| Others | 22 (11.0) |
| **Hospitalised** | 172 (82.3) |
| **Outpatient** | 37 (17.7) |

\*Includes lung, breast, and oesophageal cancer patients.

**Table 2.**
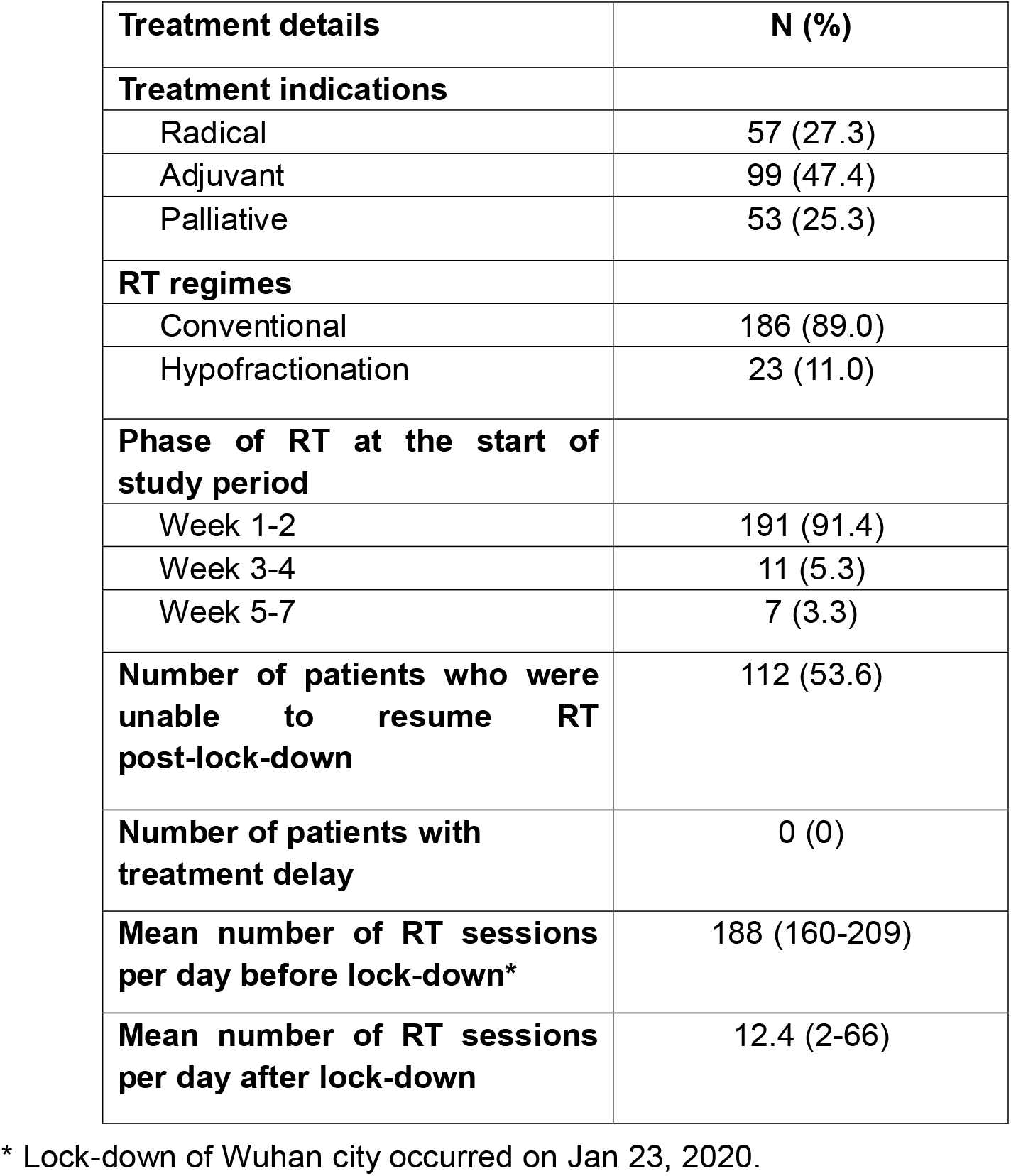
Details of the radiotherapy (RT) treatments for the 209 cancer patients.

### RT treatment details

Among 209 patients, 47.4% received adjuvant RT, while 27.3% and 25.3% of treatments were for radical and palliative intent, respectively. All patients had already commenced treatment prior to the study start-date. **Figure 3** shows the number of treated patients per day over the duration of the study. Prior to the lock-down, mean number of patients per day was 188 (range = 160-209). However, these numbers sharply declined on and immediately after the date of lock-down, and declined with subsequent weeks (mean number of patients per day = 12.4; range = 2-66).

**Figure 3.**
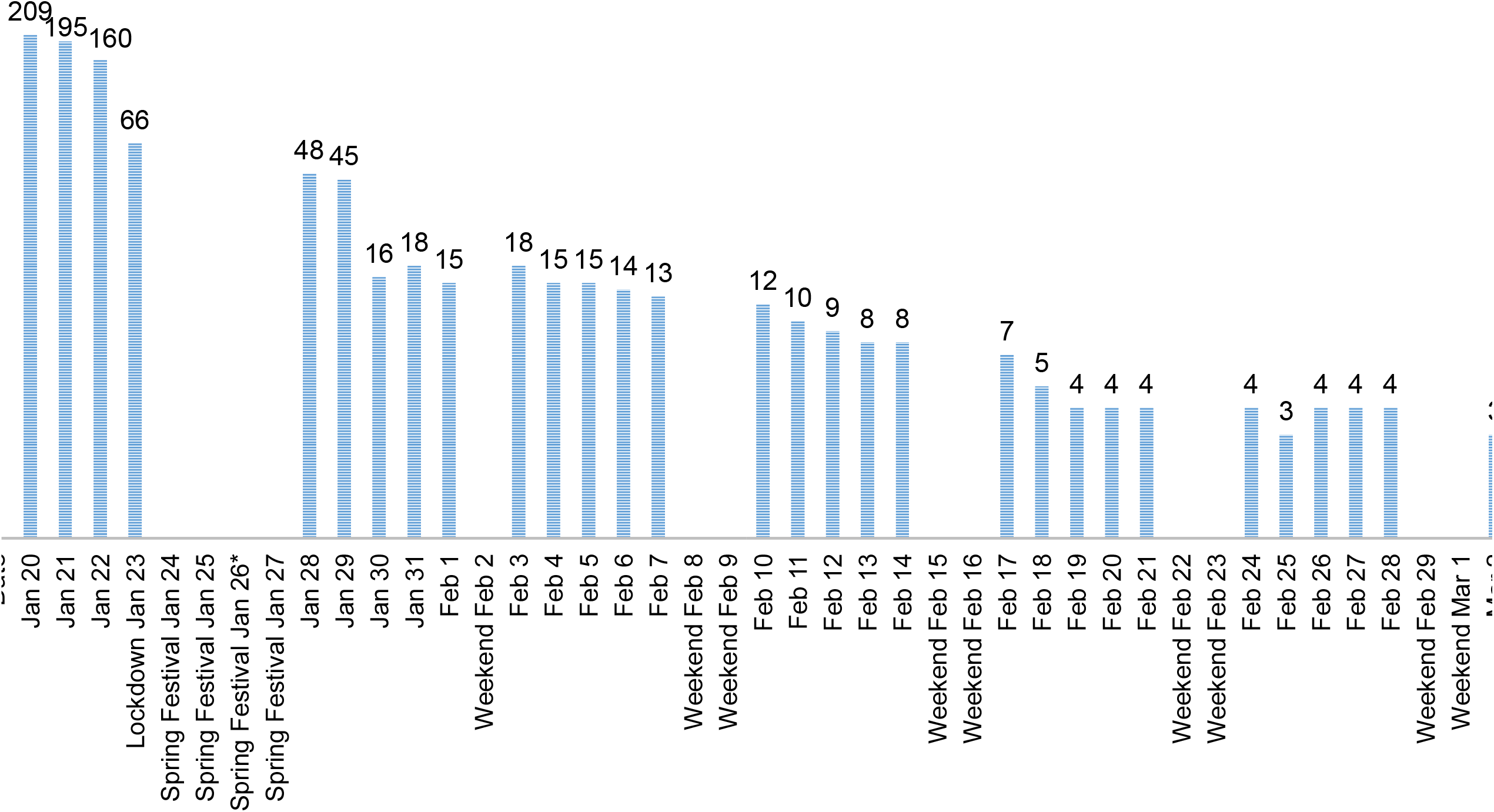
Radiotherapy case-load per day during the outbreak of COVID-19 in our department. Number of patients dropped substantially after the lock-down of the city, and declined by each week. The reduced case-load was also partly due to the restriction of admitting new cancer patients to the hospital. * Date of COVID 19 patient diagnosis

### Outcomes

As of Mar 6, 2020, we recorded only one case (0.48%) of newly-diagnosed COVID-19 pneumonia. This patient completed treatment on Jan 23, 2020, and was confirmed to have SARS-CoV-2 infection on Jan 26, 2020. Of the 209 patients, 70 were deemed to have a contact history with the COVID-19 patient; none of these patients developed clinical symptoms of COVID-19, and both CT and RT-PCR screening investigations were negative. Among them, 52 patients were not able to return for RT post-lock-down, while 18 patients continued to undergo RT without delay. Nonetheless, treatments for these patients were re-scheduled to the end of the day.

Finally, no medical staff was infected during the study period. All patients, including the COVID-19 case, remained alive as of Mar 12, 2020.

## Discussion

The outbreak of a novel SARS-CoV-2 virus has led to an unprecedented rapid spike in COVID-19 pneumonia cases globally. To date, more than 100,000 humans have been infected, and the numbers are likely to increase. Perhaps among the natural history of this virus infection, the predominant characteristic of SARS-CoV-2 is its remarkably high infectivity potential, where an infected individual can spread the virus to two to three other individuals when in close proximity.^8^ While large-scale epidemiological studies from China have suggested that young and elderly individuals are equally susceptible to COVID-19,^9^ it has also been shown that the latter subgroup, as well as patients with co-morbidities like chronic obstructive pulmonary disease and cardiac conditions are more likely to succumb to severe illness.^10^ Hence, there is a real concern about the impact of COVID-19 in patients with cancer. To date, two case series have identified that patients with cancer habour an approximately two-fold increased risk of COVID-19, and reasons underpinning these observations include older patients, immunosuppressive therapies, and recurrent hospital visitations.^5,11^ Robust infection control measures are therefore crucial to prevent cross-transmission between patients and between patient and healthcare workers within the compounds of a cancer centre.

Herein, we shared our single institution experience in the management of cancer patients undergoing RT from a cancer centre that was based in Wuhan, where the outbreak first started. Our infection control protocol was comprehensive, covering a stepwise clinical pathway to manage suspect cases (questionaire, CT, and RT-PCR assays), on-site screening, re-organisation of the RT facility, and protection of healthcare workers. The extremely low rates of infection among our patients (0.48%) and healthcare workers (0%) were a testament to the efficacy of our measures.

Notably, one patient who was treated in our RT facility became infected with SARS-CoV-2 on Jan 26, 2020 (3 days after the lock-down). We were able to establish a contact history with 70 other patients over the preceding 14-day window. Amazingly, none of the patients developed COVID-19 pneumonia on follow-up, which again highlights the effectiveness of our infection control protocol. Unfortunately, due to the lock-down, only 18 of these patients were able to resume RT, as the remaining 52 patients were not allowed to return to the city.

Some limitations of this study deserve mention. First, it is apparent that the number of patients decreased substantially following the lock-down (a 10-fold drop in case-load; **Figure 3**). While it is unknown if such a meticulous protocol would be applicable in high-volume cancer centres from other parts of the world, reduction of case-load constitutes a key step of limiting human-to-human transmission of SARS-CoV-2, and thus, patients’ treatment ought to be prioritised accordingly. Finally, it is important to assess the impact on the long-term prognosis of these cancer patients, especially for those individuals who were unable to resume their treatment, which is the undesired consequence of a lock-down and robust infection control measures.

## Conclusions

To conclude, we described infectious control measures that were undertaken to prevent human-to-human transmission among patients with cancer undergoing RT and healthcare workers. The early result of a 0.48% infection rate validates the effectiveness of our protocol. Longer-term follow-up will inform on the implications of this pandemic on the survival outcomes of cancer patients.

## Data Availability

all data available

## Author contributions

Study conception and design: CX, MC

Acquisition, analysis, or interpretation of data: CX, XW, HL, ZB, YZ

Statistical analysis: CX, JY, MC

Obtained funding: CX, MC

Administrative, technical, or material support: CX, JY, MC

Study supervision: CX, MC

Drafting of manuscript: All authors

Approval of final manuscript: All authors

## Acknowledgements

The authors would like to acknowledge all the patients, and the front-line healthcare professionals who are involved in patient care during this pandemic.

